# Dynamics of gut microbiome – mediated bile acid metabolism in progression to islet autoimmunity

**DOI:** 10.1101/2021.08.20.21262371

**Authors:** Santosh Lamichhane, Partho Sen, Alex M. Dickens, Marina Amaral Alves, Taina Härkönen, Jarno Honkanen, Tommi Vatanen, Ramnik J. Xavier, Tuulia Hyötyläinen, Mikael Knip, Matej Orešič

**Affiliations:** Turku Bioscience Centre, University of Turku and Åbo Akademi University, FI-20520 Turku, Finland; School of Medical Sciences, Örebro University, 702 81 Örebro, Sweden; Department of Chemistry, University of Turku, 20520 Turku, Finland; Walter Mors Institute of Research on Natural Products, Federal University of Rio de Janeiro, 21941-599 Rio de Janeiro, Brazil; Research Program for Clinical and Molecular Metabolism, Faculty of Medicine, University of Helsinki, Finland; The Liggins Institute, University of Auckland, Auckland, New Zealand; The Broad Institute of MIT and Harvard, Cambridge, MA, USA; School of Science and Technology, Örebro University, Örebro, Sweden; Pediatric Research Center, Children’s Hospital, University of Helsinki and Helsinki University Hospital, 00290 Helsinki, Finland

**Author notes:** Correspondence: Professor Mikael Knip, M.D., Ph.D., Pediatric Research Center, Children’s Hospital, University of Helsinki and Helsinki University Hospital, 00290 Helsinki, Finland., Professor Matej Orešič, Ph.D., Turku Bioscience Centre, University of Turku and Åbo Akademi University, FI-20520 Turku, Finland. Equal contribution (Santosh Lamichhane, Partho Sen).

**Keywords:** Bile acids, genome-scale metabolic modeling, gut microbiome, lipid metabolism, microbial metabolism, islet autoimmunity, type 1 diabetes

## Abstract

Previous studies suggest that the human gut microbiome is dysregulated in islet autoimmunity, preceding the clinical onset of type 1 diabetes (T1D). The microbiota of the gut plays an important role in the regulation of bile acid (BA) metabolism. However, not much is known about the regulation of BAs during progression to T1D. Here, we analyzed BAs in a longitudinal series of serum (n= 333) and stool (n= 304) samples, collected at 3, 6, 12, 18, 24 and 36 months of age, from children who developed a single islet autoantibody (P1Ab), multiple islet autoantibodies (P2Ab), and controls (CTRs) who remained autoantibody (AAb) negative during the follow-up. In addition, we analyzed the stool microbiome by shotgun metagenomics in a subgroup of these children (n=111). Factor analysis showed that age had the strongest impact on BA and microbiome profiles. We found that, at an early age, the systemic BA (including taurine and glycine conjugates) and microbial secondary BA pathways were altered in the P2Ab group as compared to the P1Ab or CTR groups. Our findings thus suggest that dysregulated BA metabolism in early life may contribute to the risk and pathogenesis of T1D.

## Introduction

Bile acids (BAs) are amphiphilic molecules that are crucial physiological agents for facilitating the absorption of lipids in the small intestine. BAs are produced from cholesterol in the liver. Primary BAs such as cholic acid (CA) and chenodeoxycholic acid (CDCA) are conjugated with either glycine or taurine in the hepatocytes (de Aguiar Vallim *et al*, 2013). Gut microbes transform primary BAs to secondary BAs in the intestine (Ramírez-Pérez *et al*, 2017). Most of these BAs are re-absorbed back to the liver, while approximately 5% of the total BA pool is excreted *via* feces. Under normal physiological conditions, a small fraction (about 10%) of the BAs are re-circulated and enter the systemic (enterohepatic) circulation, where they act as ligands for receptors in the peripheral tissues, including farnesoid X receptor (FXR) and the membrane receptor known as Takeda G protein-coupled membrane receptor (TGR5) (Chiang, 2013; Makishima *et al*, 1999; Wang *et al*, 1999). FXR and TGR5 signaling plays a critical role in regulating the systemic lipid, glucose, and energy homeostasis (Jiang *et al*, 2015; Shapiro *et al*, 2018). Dysregulation of systemic BA metabolism has been linked to multiple diseases, including fatty liver disease, cardiovascular disease, and type 2 diabetes (Chan & Wong, 2019; Shapiro *et al*., 2018; Zheng *et al*, 2021). Thus, the gut-microbiome-BA axis is increasingly recognized as a therapeutic target for treating metabolic and immune disorders (Chan & Wong, 2019; Gu *et al*, 2017; Targher *et al*, 2021).

Previous metabolome and gut microbiome studies suggest that children who progress to islet autoimmunity and type 1 diabetes (T1D) later in life, are characterized by dysregulation of lipid metabolism (Lamichhane *et al*, 2018; Li *et al*, 2020; Oresic *et al*, 2008; Sen *et al*, 2020) and gut microbiota (Kostic *et al*, 2015; Siljander *et al*, 2019; Vatanen *et al*, 2016), suggesting that there is an interplay between host metabolism, the immune system, and the gut microbiome during early T1D pathogenesis. However, our understanding of both microbial and host regulation of BA pathways in the development of islet autoimmunity is still scarce (Kostic *et al*., 2015).

Herein, we set out to investigate how microbial BA pathways are regulated in children who develop islet autoimmunity. We analyzed BAs and subject-matched microbiome profiles in a prospective series of samples, which included children who developed multiple autoantibodies (P2Ab) during follow-up and are thus at high risk of progression to T1D later in life (Ziegler *et al*, 2013), and those children who developed only one islet autoantibody (P1Ab) but did not progress to T1D during follow-up. We also included control children (CTRs) who remained islet autoantibody (AAb) negative during follow-up.

## Results

### Prospective study of bile acids and gut microbiome in children at risk for T1D

We analyzed BAs prospectively in subject-matched stool (n= 304) and serum (n= 333) samples from three study groups: P1Ab (n=23), P2Ab (n=13), and CTR (n=38) (**Figure 1**). From each child, we analyzed stool and serum samples at six different time-points corresponding to the ages of 3, 6, 12, 18, 24, and 36 months. A total of 33 BAs, including both primary (glycine/taurine conjugates) and secondary BAs were assayed. Previously-published stool shotgun metagenomics data (whole-genome shotgun sequencing (WGSS)) from a subset of children (n=111 stool samples in total) (Kostic *et al*., 2015) were included in the study (**Figure1**).

**Figure 1.**
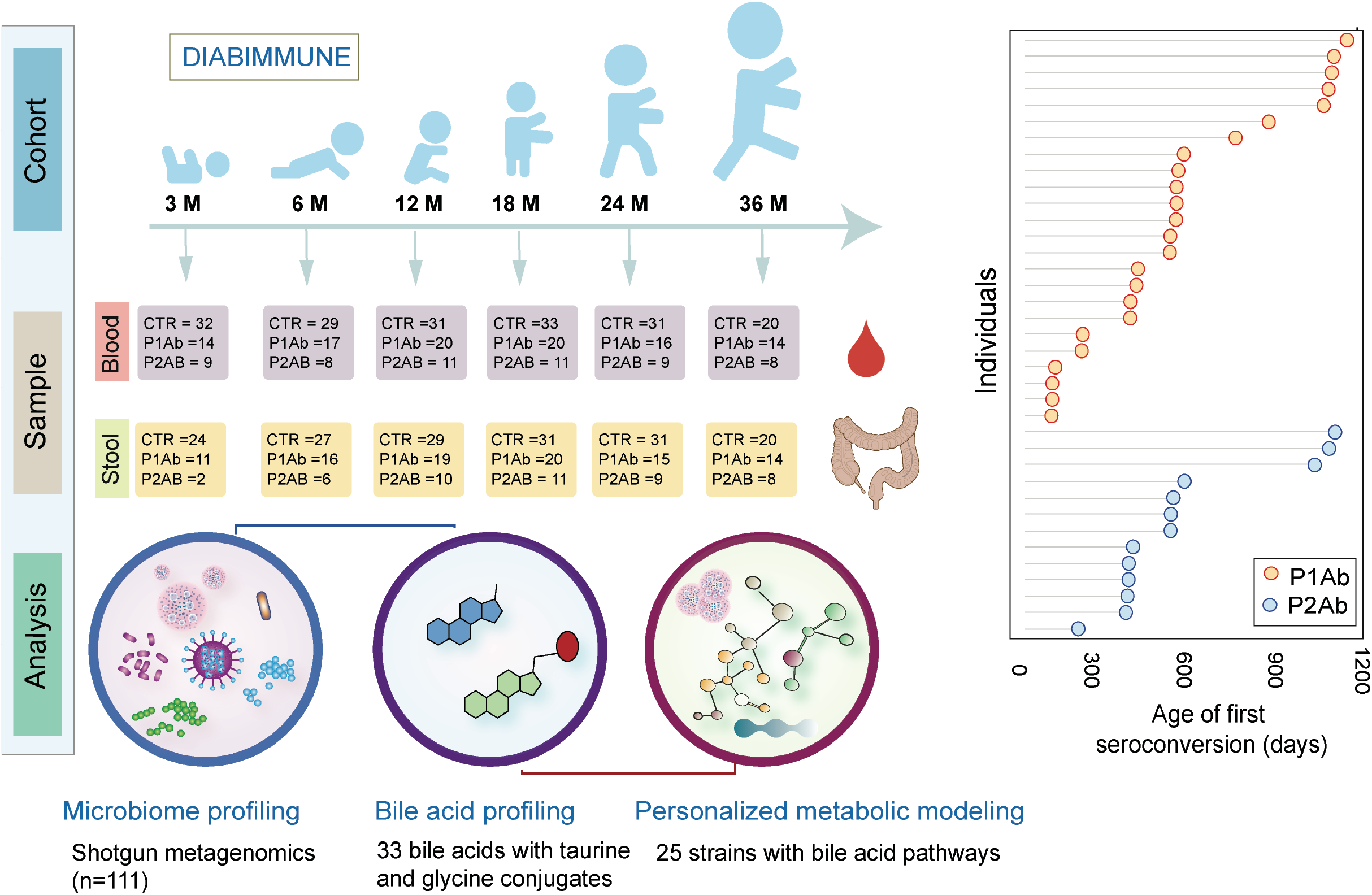
Outlines of analytical study flow. This illustrates the number of serum and stool samples collected for targeted BA measurement at each time-point. The samples were stratified into P1Ab, P2Ab and CTR groups. Moreover, this shows the age at seroconversion among the children taking part in this study.

### Age-related changes in bile acid and microbiome profiles

In order to determine the contributions of various factors to the BA profiles, multivariable associations were tested for, applying linear models using co-variates including age, sex, and case status (P1Ab, P2Ab, or CTR), taking into account random effects within an individual sample/subject. Age showed the strongest impact (23 stool and 21 serum BAs at p < 0.05; **Figure 2A**), while five stool and one serum BA were different across case groups and one stool and four serum BA were different between sexes (**Tables S1** and **S2**). Primary BAs, including cholic acid (CA) and chenodeoxycholic acid (CDCA), were decreased both in stool and in circulation with increasing age (**Figure 2A**). A similar trend was seen for deoxycholic acid (DCA), a secondary BA. Low levels of other secondary BAs (including their taurine and glycine conjugates) were observed during early infancy (3 and 6 months), which steadily increased at/after the first year of life (12 and 18 months) and remained stable at later ages (24 and 36 months) (**Figure 2A**).

**Figure 2.**
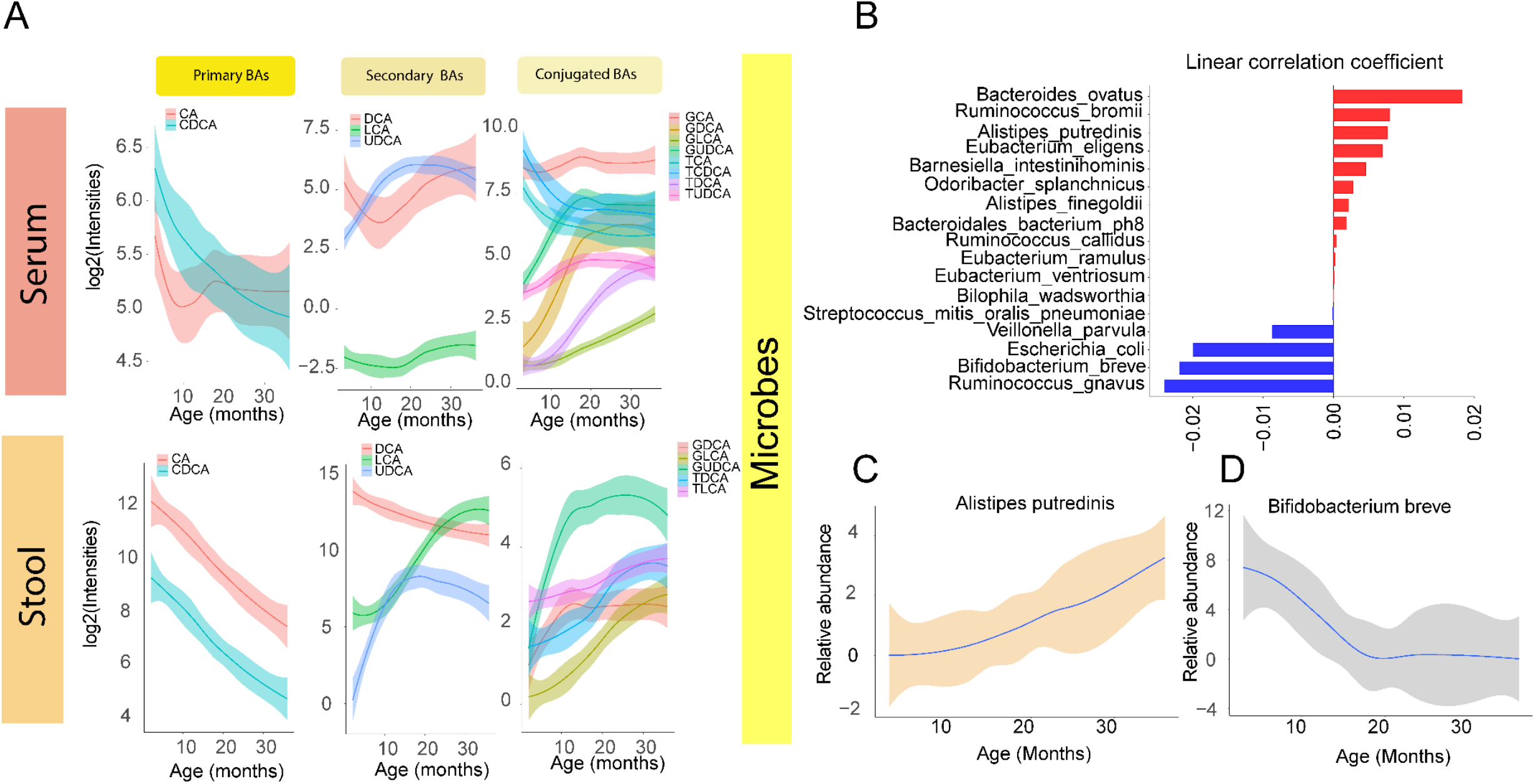
Age-related changes in the bile acid and microbiome. **(A)** The loess curve plot of BAs over time (3, 6, 12, 18, and 24) between stool and serum samples. This panel plots, separately, representative primary, secondary and conjugated BAs that changed significantly over time (p < 0.05). **(B)** Bar plots showing correlation coefficients for the association between age and different microbes. Red represents inverse correlations while blue represents positive correlations as obtained by multivariate linear regression using MaAsLin2 R package. **(C-D)** The loess curve plot of selected microbes over time.

Gut microbial profiles follow the dynamic BA trajectories (**Figure 2A-B, Table S3**). Intuitively, age was the strongest factor associated with the composition of the infant gut microbiome (**Figure S1**). Several microbial species, at the strain level, were distinctively associated (n = 17, p < 0.05) with age (**Figure 2B, Figure S1**); dominated by *Ruminococcus, Alistipes*, and *Eubacterium* species (spp.) which showed an increasing trend with age (**Figure 2D**). However, this did not stabilize at 36 months of age. On the other hand, the abundances of six out of 17 microbes, including *Bifidobacterium breve*, (**Figure 2D**) were initially decreased during 3 to 12 months of infancy and stabilized at 24 and 36 months of age.

### Alteration of the gut microbiome impacts bile acid metabolism in progression to islet autoimmunity

Differential analysis showed that 41 microbial strains were altered (analysis of covariance, ANCOVA; Tukey’s honest significant differences (HSD), p.adjusted < 0.05) between the study groups (P1Ab and/or P2Ab and/or CTRs), at at least one time-point (**Figure 3A**). Of note, 25 of 41 microbes were known to exhibit BA metabolic pathways annotated by the Assembly of Gut Organisms through Reconstruction and Analysis (AGORA) compendium (Heinken *et al*, 2019; Magnusdottir *et al*, 2017; Noronha *et al*, 2019). Among these, several strains of *Clostridium, Lachnospiraceae, Ruminococcus* and *Alistipes* were predominant and were altered between the P1Ab and P2Ab groups (**Figure 3A**). Lower abundances of *Clostridium and Lachnospiraceae* strains and increased abundances of *Ruminococcus* strains were apparent between P1Ab *vs*. P2Ab at 18 months and/or 24 months of age.

**Figure 3.**
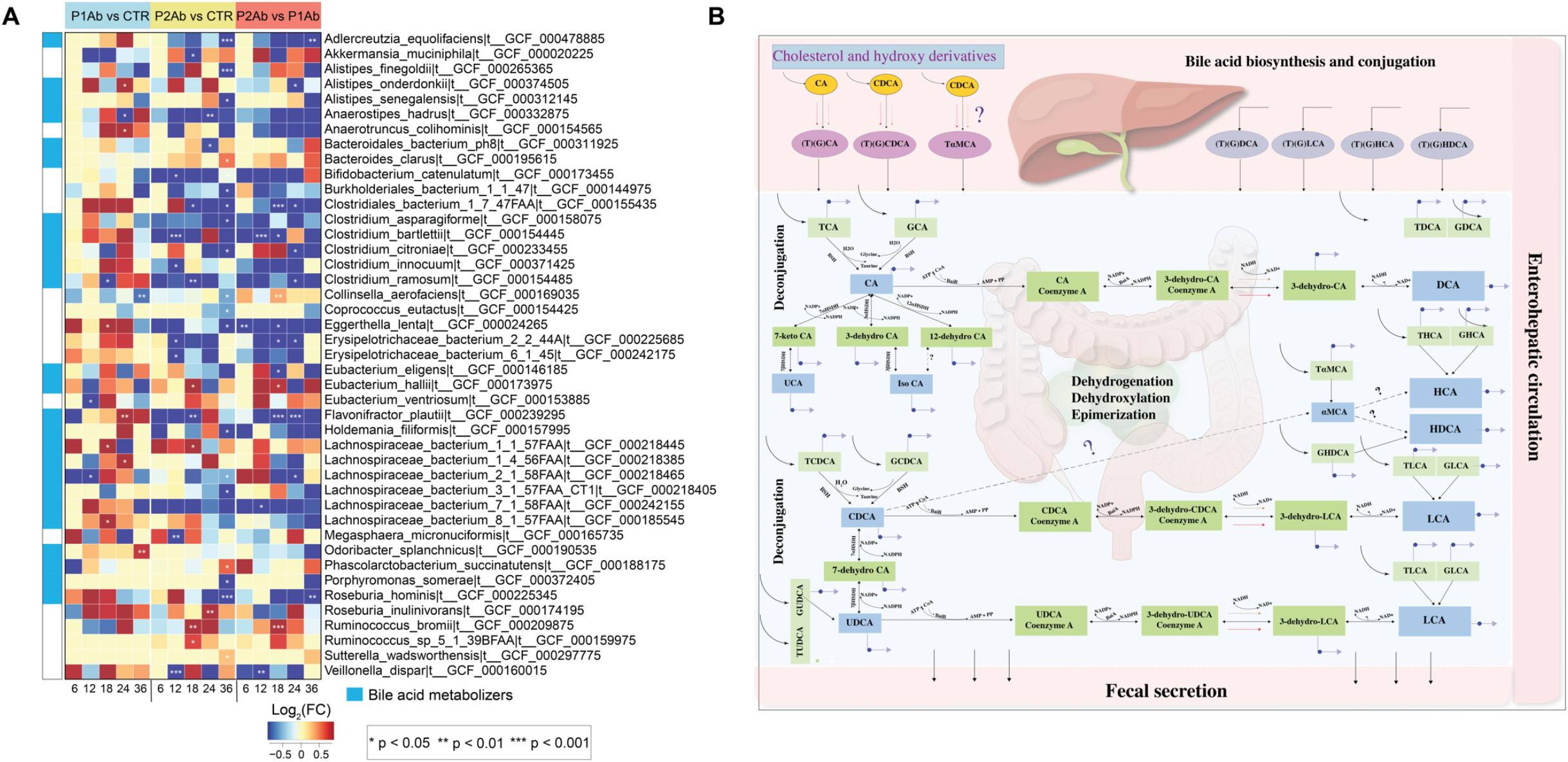
Microbial strains in progression to islet autoimmunity. **(A)** A heatmap showing the log2 fold changes (FCs) in the strain-level abundances of the gut microbes in P1Ab *vs*. CTRs, P2Ab *vs*. CTRs and P2Ab *vs*. P1Ab groups at 6, 12, 18, 24 and 36 months of the follow-up. Red, blue and yellow color denotes increase, decrease and no change in the abundances between the differential conditions, respectively. Statistical significance was assessed by ANCOVA adjusted for ‘age’ as a covariate, and Tukey’s HSD (p.adjusted < 0.05). Microbes with BA pathways (annotated by the AGORA compendium) are marked with light blue color. **(B)** An illustration of BA metabolism and related pathways in the humans.

Pairwise metabolic modeling between microbes revealed that most of these microbes exhibit amensal (50%), parasitic (20%) and commensal (15%) relationships for cross-feeding and biotransformation of BAs in the human gut (**Figure S2**). Several strains such as *Roseburia hominis A2-183, Roseburia inulinivorans DSM 16841, Ruminococcus bromii L2-63, Lachnospiraceae bacterium sp. 5_1_63FAA* can only produce secondary BAs, whilst *Anaerostipes hadrus DSM 3319, Bifidobacterium catenulatum DSM 16992, Bifidobacterium coryneforme DSM 20216, Clostridium bartlettii DSM 16795, Clostridium innocuum 2959, Collinsella aerofaciens ATCC 25986, Coprococcus eutactus ATCC 27759, Eggerthella lenta DSM 2243, Erysipelotrichaceae bacterium 5_2_54FAA, Erysipelotrichaceae bacterium sp. 3_1_53, Eubacterium eligens ATCC 27750, Eubacterium hallii DSM 3353* can interact with at least 10 other microbial strains to carry out BA biotransformation (**Figure S2**). Interestingly, *Alistipes spp*. co-exhibited different kinds of microbial interaction, including amensalism, parasitism and commensalism for BA biotransformation (**Figure S2**). The BA pathways exhibited by these microbes include ten different reaction classes that can carry out deconjugation, dehydrogenation, dehydroxylation and epimerization of BAs in the human gut **(Figures 3B** and **S3**).

### Regulation of secondary bile acid pathways before the emergence of islet autoantibodies

In order to understand the interplay between the gut microbiome and BA biotransformation in the progression to islet autoimmunity, we developed personalized community microbiota models for each child. The community microbiota model comprises 25 abundant microbial strains and their BA reactions (**Figure 3A, Figure S3**).

The community microbiota modeling suggested that the total BA reaction abundances were markedly decreased (Tukey’s HSD, p.adjusted < 0.05) in the P2Ab *vs*. P1Ab groups at 6 and 12 months of age, *i*.*e*., before the median age of seroconversion (**Figure 4A**). Moreover, at this age, the predicted abundances of bile salt hydrolases (BSH) reaction(s) decreased in the P2Ab *vs*. P1Ab group. However, the abundances of these reactions peaked at 24 months of age (post seroconversion) (**Figure 4B**). At this age, several reactions in the alpha/beta dehydroxylation pathway (12-alpha-, 3-alpha-, 7-beta-hydroxysteroid dehydrogenases and cholate ligase) showed decreased abundances in the P2Ab group (*vs*. P1Ab). The abundances of these reactions peaked at 18 months of age (median age of seroconversion). 7-alpha/beta hydroxylation pathways aid in the production of secondary BAs (*e*.*g*., DCA, HCA, HDCA and LCA) from primary BAs (*e*.*g*., CA and CDCA), respectively (**Figures 3B** and **4B**).

**Figure 4.**
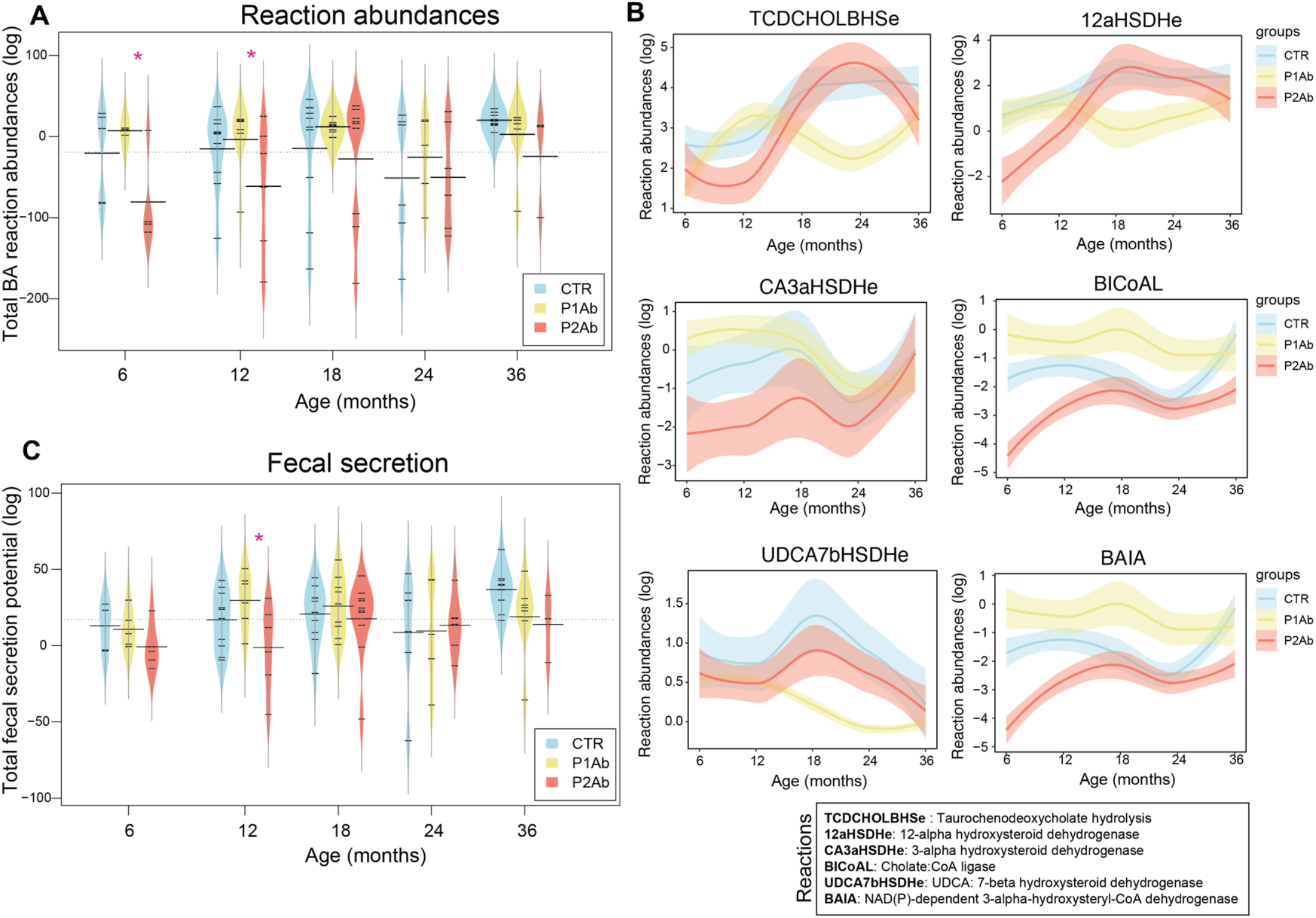
Regulation of bile acid reactions in progression to islet autoimmunity. **(A**,**C)** Beanplots showing the levels of total BA reaction abundances and the total fecal secretion potentials (FSPs) predicted by the community microbiota models in CTRs, P1Ab and P2Ab groups at 6, 12, 18, 24 and 36 months of the follow-up. The black dotted line denotes the mean of the population. The black dashes in the bean plots represent the group mean. ‘*’ denotes significant differences (ANCOVA + Tukey’s HSD, p.adjusted < 0.05). **(B)** Locally Weighted Scatterplot Smoothing (LOWESS) plot showing the longitudinal trend of an individual BA reaction abundance in the CTRs (light blue), P1Ab (yellow) and P2Ab (orange) groups during the follow-up. The shaded area around the curves depicts the 95% confidence interval.

The community modeling also suggested that the total fecal secretion potential (FSP) of secondary BAs was significantly decreased (Tukey’s HSD, p.adjusted < 0.05) in P2Ab (*vs*. P1Ab) group at 12 months of age (**Figure 4C**). Taken together, secondary BA production seems to be decreased in the P2Ab group as compared to P1Ab and/or CTR group(s) with the emergence of islet autoantibodies.

### Targeted measurements of BAs revealed a decrease in secondary BA levels in progression to islet autoimmunity

Next, we sought to determine specific BA concentration differences between the three study groups in the longitudinal series of stool and serum samples. We detected several differences in BA concentrations between the study groups. In particular, taurine and glycine conjugates of secondary BAs (*e*.*g*., GHDCA, GUDCA, TUDCA, THDCA) were decreased in the P2Ab group at 6 and 12 months of age in stool samples, as compared to the CTR or the P1Ab groups (**Figure 5A**). The levels of GLCA, wMCA and LCA were higher in P2Ab than in P1Ab at 18 months of age. No clear differences were seen for the primary BAs, which further suggests that subsequent steps of BA dehydroxylation and secondary BA production were (dys)regulated in the P2Ab group (**Figure 3B**). However, these differences were less pronounced in the serum samples as compared to the stool samples (**Figure 5**). Moreover, there was no persistent longitudinal trend with respect to the differences in BA levels between the groups throughout follow-up, with the exception of TwMCA which was persistently decreased in P2Ab *vs*. P1Ab and CTR groups.

**Figure 5.**
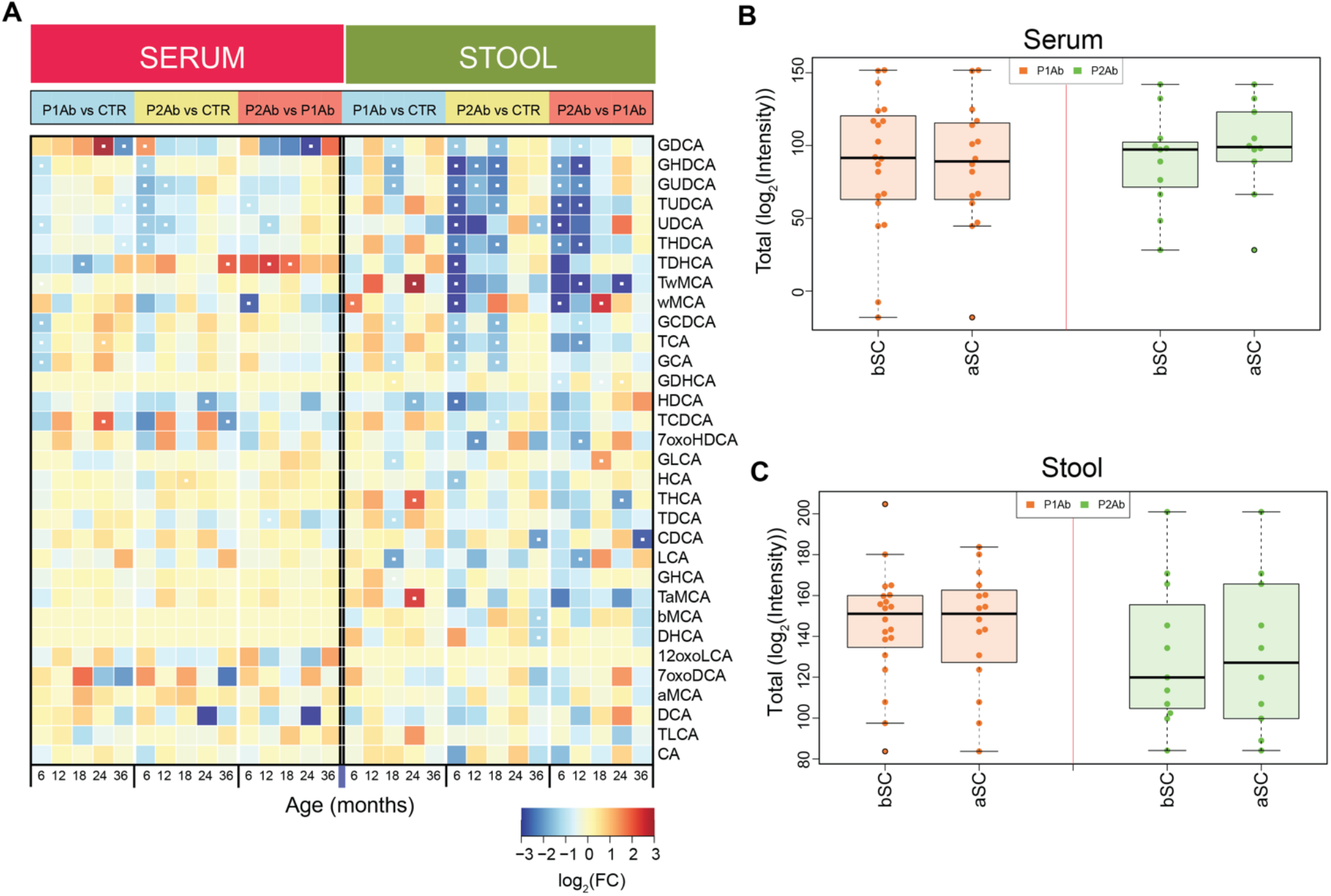
Systemic alterations in bile acid profiles in progression to islet autoimmunity. (**A)** A heatmap showing the log2 fold changes (FCs) in BA profiles in P1Ab *vs*. CTRs, P2Ab *vs*. CTRs and P2Ab *vs*. P1Ab groups at 6, 12, 18, 24 and 36 months of follow-up. Red, blue and yellow color denotes increase, decrease and no change in the intensities of BAs between the differential conditions, respectively. Statistical significance was estimated by ANCOVA adjusted for ‘age’ as a covariate, and Tukey’s HSD (p.adjusted < 0.05). **(B-C)** Boxplot showing the total intensities of BAs before and after seroconversion in P1Ab and P2Ab groups. Statistical significance was estimated by a paired t-test (p < 0.1).

Next, we compared the total stool and serum BA concentrations in the P1Ab and P2Ab groups in samples obtained before and after the appearance of the first islet autoantibody, respectively. Pairwise analysis (paired t-test, p < 0.05) revealed that, before the emergence of islet autoantibodies, the total BA pool, both in stool and serum, was lower in the P2Ab group (**Figure 5B-C**).

### Association between BAs, gut microbes and their exchange potentials shows unique patterns of regulation in the P2Ab group

At 12 months of age, the UDCA exchange potential of the gut microbiota was positively associated with several species of *Alistipes* and *Eubacterium*, particularly *Eubacterium ventriosum ATCC 27560* (p.adjusted < 0.05). Furthermore, it was positively associated with the GLCA levels (a downstream product) in stool (**Figure 6A)**. At 18 months, the UDCA exchange potentials were strongly associated with *Roseburia hominis A2-183 and Roseburia inulinivorans DSM 16841ı (*p.adjusted < 0.05), whilst inversely associated with the conjugated and/or unconjugated secondary BAs in the stool samples (**Figure 6B)**. *Clostridium bartlettii DSM 16795* was positively associated with the exchange potential of T (DCA) at 12 months, whereas there was an inverse association at 18 months. Several species of *Alistipes* were inversely associated (p.adjusted < 0.05) with TLCA (p.adjusted < 0.05) (**Figure 6A-B**).

**Figure 6.**
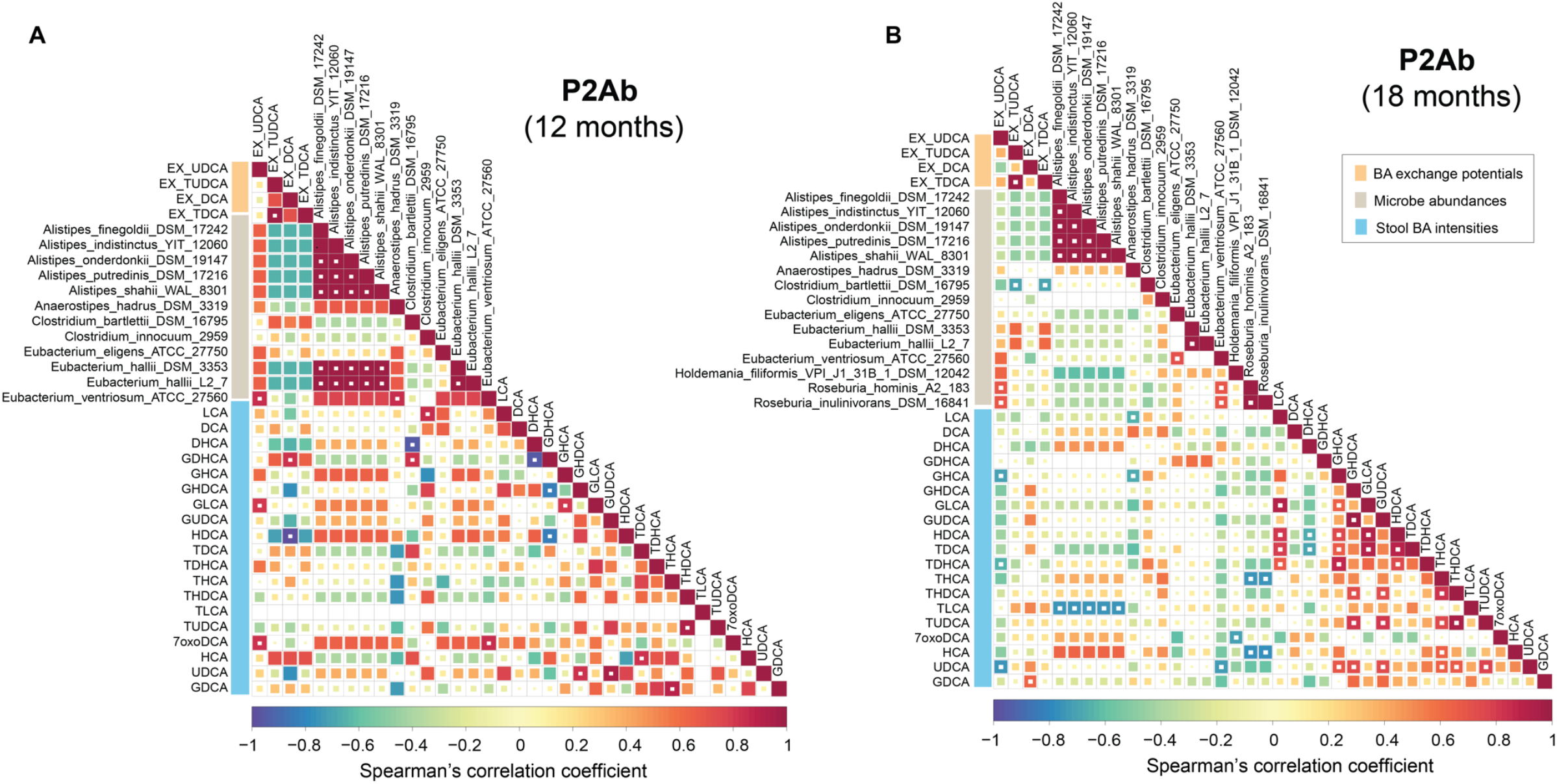
Cross-correlation between the gut microbiome, BA exchange potentials and their systemic (stool) levels in progression to islet autoimmunity. Correlation plots showing bivariate Spearman’s correlations between the gut microbiome (metagenomics) and/or BA exchange potential (microbiota community model) and/or level of BA (lipidomics) in the stool samples of P2Ab (n=111) group at 12 and 18 months. Red, blue and white/yellow color represents positive, negative and no correlation respectively. The white ‘dot’ depicts that the correlation is statistically significant (p.adjusted < 0.05).

## Discussion

By combining targeted metabolomics and metagenomics data, we show that host-microbial BA co-metabolism is dysregulated in the progression to islet autoimmunity and overt T1D. Our findings suggest that children who progress to multiple islet autoantibodies (P2Ab) during follow-up, and are thus at high risk of developing T1D later in life (Ziegler *et al*., 2013), have distinct and persistently-altered systemic BA concentrations and species abundances in the gut microbiome, as compared to those children who develop, at most, a single islet autoantibody (P1Ab), or those who remained negative for islet antibodies (CTRs) during the follow-up.

Our results reveal that children who progressed to multiple islet autoantibodies later in life had decreased concentration of conjugated BAs (GHDCA, GUDCA, TUDCA, and THDCA) in early life. In line with this, tauroursodeoxycholic acid (TUDCA), a conjugated secondary BA, was observed to reduce the incidence of diabetes development by improving the glucose utilization and metabolism in the streptozotocin administered C57BL/6 mice (Bronczek *et al*, 2019). Our personalized community metabolic modeling of the gut microbiota identified specific differences in the BA pathways of P2Ab *vs*. P1Ab or CTR groups. Several intermediary reactions of 7-alpha/beta hydroxylation and bile salt hydrolase (BSH) pathways were altered at or before the age of seroconversion. Alteration of relative abundances of BSH levels are associated with the occurrence and development of various diseases in humans (Song *et al*, 2019). Here, we revealed that the activity of BSH may be related to the development of islet autoimmunity and risk of clinical T1D. Furthermore, our results show that in the P2Ab (*vs*. P1Ab) group, reaction abundances of 7-alpha/beta hydroxylation pathway remained lower at 6 and 12 months of age, which gradually increased at later time-points. BSH pathways are key gatekeepers of BA transformation in the gut (Foley *et al*, 2019). We found stool concentration of secondary BAs, particularly UDCA, DCA, HDCA and their glycine and/or taurine conjugates, were downregulated in P2Ab *vs*. P1Ab and/or CTR groups. Decreased fecal secretion potentials (FSPs) of the secondary BAs in the P2Ab group during early life further support the view that a decrease in secondary BA levels at or before the age of seroconversion might occur due to a decrease in the metabolic potential of the microbiota-encoded 7-alpha/beta hydroxylation pathway, which aids in the transformation of secondary BAs.

We also found that BA concentrations are strongly dependent on the age of the children. In agreement with previous findings (Backhed *et al*, 2015; Milani *et al*, 2017), we observed that the abundances of gut microbes (with the exception of a few strains) gradually increased with the age of the children. Interestingly, many of these microbes are involved in the biotransformation of BAs (Heinken *et al*., 2019; Magnusdottir *et al*., 2017). Recently, a study characterized the age-dependent gut microbial and metabolic changes in the murine gastrointestinal tract (van Best *et al*, 2020), where BAs were identified as a major driver for the early maturation of the gut microbiome.

We acknowledge some limitations of our study, such as the relatively small sample size, as well as the effect of a lack of full, systematic understanding of the impact of confounding lifestyle factors (*e*.*g*. diet) and these factors’ relationship with BA host-co-metabolism. Nevertheless, we report BA changes in a longitudinal setting, defining the time-course of changes in BA metabolism with respect to the onset of islet autoimmunity. It is, however, clear that these findings need to be investigated and replicated in other, larger studies and in heterogeneous populations.

In summary, our findings suggest that dysregulated BA metabolism in early life may contribute to the risk and pathogenesis of T1D. BA metabolism may also be an underlying link between the gut microbiome and host (lipid) metabolism during the period preceding seroconversion to positivity for islet autoantibodies and overt T1D.

## Methods

### Clinical study setting

The DIABIMMUNE study recruited 832 families in Finland (Espoo), Estonia (Tartu), and Russia (Petrozavodsk) with infants carrying HLA alleles that conferred risk for autoimmunity. The subjects involved in the current study were chosen from the subset (n = 74) of international DIABIMMUNE study children who progressed to at least a single AAb (P1Ab, n = 23), who progressed to multiple islet AAb (P2Ab, n = 13), and controls (CTRs, n = 38), *i*.*e*. the children who remained islet AAb-negative during the follow-up in a longitudinal series of samples collected at 3, 6, 12, 18, 24 and 36 months from each child (Kostic *et al*. 2015). The study groups were matched for HLA-associated diabetes risk, sex, country and period of birth.

This study was conducted according to the guidelines in the Declaration of Helsinki. The Ethics Committee of the Hospital District of Helsinki and Uusimaa approved the study protocol. All families provided written informed consent prior to sample collection.

### Quantification of bile acids

The bile acids were measured in serum and fecal sample as described previously (Jäntti *et al*, 2014; Salihović *et al*, 2020). Briefly, 20 μL of serum, or fecal homogenate (prepared by adding 1:20 (m/v) ultrapure water to 50 mg of feces) was filtered through a Ostro Protein Precipitation and Phospholipid Removal 96-well plate (Waters Corporation, Milford, USA), using 100 μL of cold methanol contemning the internal standard mixtures (LCA-d4, TCA-d4, GUDCA-d4, GCA-d4, CA-d4, UDCA-d4, GCDCA-d4, CDCA-d4, DCA-d4, GLCA-d4). The eluent was collected and evaporated to dryness and the residue was re-suspended in 20 μL of a 40:60 MeOH: H2O v/v mixture. The analyses were performed on an ACQUITY HSS T3 (2.1×100 mm, 1.8 μm) column, Waters (Milford), coupled to a triple quadrupole mass spectrometer (Waters Corporation, Milford, USA) with an atmospheric electrospray interface operating in negative ion mode. Separation was performed using gradient elution with 0.1 % formic acid in water (v/v) (A) and 0.1 % formic acid in acetonitrile:methanol (3:1, v/v) (B) at a flow rate of 0.5 mL/ min. Gradient program was 0 min 15 % B, 0-1 min; 30 % B, 1-16 min; 16-18 min; 70 % B, 18–23 min 100 % B, and equilibrium time between runs was 7 min. The injection volume was 5 μL and the column was kept at 35 °C. An external calibration with nine calibration points (0.0025–600 ng/mL) was carried out for use in quantitation.

### Analysis of islet autoantibodies

Four diabetes-associated autoantibodies were analyzed from each serum sample with specific radiobinding assays: insulin autoantibodies (IAA), glutamic acid decarboxylase antibodies (GADA), islet antigen-2 antibodies (IA-2A), and zinc transporter 8 antibodies (ZnT8A) as described previously (Knip *et al*, 2010). Islet cell antibodies (ICA) were analyzed with immunofluoresence in those subjects who tested positive for at least one of the autoantibodies. The cut-off values were based on the 99th percentile in non-diabetic children and were 2.80 relative units (RUs) for IAA, 5.36 RU for GADA, 0.78 RU for IA-2A and 0.61 RU for ZnT8A.

### Taxonomic / phylogenetic profiling and metagenomic analysis

Raw metagenomic sequencing data was retrieved from (https://diabimmune.broadinstitute.org/) (NCBI BioProject ID: PRJNA231909) (Kostic *et al*., 2015). Stool (n = 111) samples were common between the published metagenomics data (Kostic *et al*., 2015), and the stool BAs measured in the present study. Metagenomic data form the matched samples (n=111) were considered for further analysis.

As stated in (Kostic *et al*., 2015), host genome-contaminated reads and low-quality reads are already removed from the raw sequencing data using kneadData v0.4. Taxonomic microbiome profiles were determined using MetaPhlAn2 (Truong *et al*, 2015) using default parameters.

### Genome-scale community modeling of human gut microbiota

Previously, genome-scale metabolic modeling (GSMM) using an Assembly of Gut Organisms through Reconstruction and Analysis (AGORA) approach has been used to elucidate the role of gut microbiota in BA biotransformation in humans (Heinken *et al*., 2019). In addition, it has been used to estimate the metabolic capabilities of gut microbes and related pathways under different biological conditions (Magnusdottir *et al*., 2017).

We used GSMM to model the dynamics of BA metabolism aided by human gut microbiota under various conditions. In order to reduce the complexity of community modeling, we included genome-scale metabolic models (GEMs) of 25 abundant gut microbes (strains) that have BA metabolic pathways, and were significantly (ANCOVA; Tukey’s HSD p.adjusted < 0.05) altered between the study groups (P1Ab, P2Ab and CTRs), at at least one time-point (**Figure 3A**). All the microbial-GEMs obtained were retrieved from the *‘AGORA_BA’* compendium (v1.03) (Heinken *et al*., 2019; Magnusdottir *et al*., 2017), stored at the Virtual Metabolic Human Database (VMH) (Noronha *et al*., 2019) and assessed for further analysis.

Next, we developed personalized community models for each individual by contextualizing the community microbiota model with the metagenomic abundances of the microbes estimated for each individual/sample. A detailed protocol for integration of metagenomic abundances into a community microbiota model has been described elsewhere (Baldini *et al*, 2019; Heinken *et al*., 2019; Magnusdottir *et al*., 2017). We performed three types of analysis, (1) Modeling the primary growth of the individual microbe based on Average European Diet (AED) (Magnusdottir *et al*., 2017; Noronha *et al*., 2019), (2) Microbe-microbe pairing (co-growth) of 25 abundant microbial strains, to determine the joint ability of the microbes for BA biotransformation, and (3) Personalized community modeling at an individual sample level using metagenomics data. The simulation results were divided for three different study groups (P1Ab, P2Ab and CTRs). GSMM was performed using the COBRA Toolbox (Heirendt *et al*, 2017) and the Microbiome Modeling Toolbox (MMT) (Baldini *et al*., 2019) deployed in MATLAB *Inc*., version R2017a.

The 25 abundant microbial-GEMs were coupled into a community microbiota model using the *‘joinModelsPairwiseFromList’* function implemented in the MMT (Baldini *et al*., 2019). A common luminal compartment ‘[u]’ was introduced that enabled metabolite exchanges and cross-feeding between the microbes. The composition of AED was retrieved from (VMH). The intestinal luminal uptake rates were constrained by the dietary fluxes using *‘useDiet’* function. The paired models were allowed to exchange conjugated bile acids (BAs), while the uptake of other metabolites was limited. Different types of metabolic interactions were estimated by the *‘simulatePairwiseInteractions’* function deployed in MMT with a minimum growth rate difference of 10% between the microbes. Each microbial model (GEM) was allowed to grow on its own, and as a pair under anaerobic conditions. Flux Variability Analysis (FVA) using *‘fastFVA’* function was performed to assess maximum and minimum fluxes incurred by the BA reactions in a paired model. The paired models that carried fluxes through the BA exchange reactions were selected and grouped by their genus (**Figure S2**).

Several metabolic reconstruction/models such as Recon3D (Brunk *et al*, 2018), the small intestinal epithelial cells (sIECs) (Sahoo & Thiele, 2013) model, and the VMH database and bibliographic references were mined, and putative BA transporters in the human gut were identified. The BA transporters and exchange reactions were added. Recon3D as a host model was coupled with the community microbiota model using the *‘createMultipleSpeciesModel’* function coded in MMT, subsequently the flux coupling constraints were added. A compartment ‘[b]’ for body fluids was introduced. Sanity checks were performed using the COBRA Toolbox. All community microbiota models were able to carry out basic metabolic tasks, including exchange and transport of BAs. The average metabolic reactions and metabolites of a microbiota community model was 15,800 and 13,900 respectively.

The fecal secretion potential (FSP) of a BA reaction is given by FSP_ij_ = A_i_ × v_j_ (eq.1), where ‘FSP_ij_’ denotes the estimated potential of ‘j^th^’ BA in ‘i^th^’ species. A and v represent the relative abundance of a species and absolute flux potential (mmol/gDw/day), respectively (Heinken *et al*., 2019; Kumar *et al*, 2018). FSP determines the metabolic efficiency of a particular reaction, under a specified condition. The total FSP determines the metabolic capability/potential of the gut microbes in a community to perform a particular task. Likewise, BA reaction abundances in a community model was estimated by the *‘calculateReactionAbundance’* function coded in MMT (Heinken *et al*., 2019; Noronha *et al*., 2019).

### Statistical analysis

The R statistical programming language (v4.0.4) and MATLAB *Inc*., (vR2017a) was used for data analysis. The *‘Heatmap*.*2’, ‘boxplot’, ‘beanplot’*, ‘*gplot*’, and ‘*ggplot2*’ R libraries / packages were used for data visualization.

#### Impact of clinical / demographic factors on stool microbiome

The effect of different factors such as age, sex, presence of antibodies, age of T1D onset, duration of breast feeding, HLA-risk class on the microbiome abundances were evaluated for each sample, and the % of explained variance (EV) was estimated. The data were log2-transformed, centered to zero mean and unit variance (autoscaled). The relative contribution of each factor to the total variance in the dataset was estimated by fitting a linear regression model, where the normalized abundances of the microbes were regressed to the factor of interest, and thereby median marginal coefficients (R^2^) were estimated. This analysis was performed using the *‘Scater’* package in R (v4.0.4). Age was found to be a confounding factor (>10% EV).

#### Differential abundant analysis of the microbiome and BAs across multiple conditions

The metagenomic and BA data were log2-transformed. By combining analysis of covariance (ANCOVA) adjusted for ‘age’ as a covariate, and posthoc Tukey’s HSD test (controls for Type I family-wise error rates at p < 0.05), we were able to identify differentially abundant microbes (p.adjusted < 0.05) between a paired condition (*e*.*g*. P2Ab *vs*. CTRs). This analysis was performed by *‘aov’* and *‘TukeyHSD’* functions deployed in the *‘stats’* package (R v4.0.4). Multivariable associations using linear models were performed using ‘MaAsLin2’ R package (Mallick *et al*, 2021). The locally-weighted regression plot was made using smoothing interpolation function loess available from ggplot2 package in R. Loess regression was performed using *‘loess’* function deployed in the *‘stats’* package (R v4.0.4).

#### Bivariate correlation analysis

*‘RcmdrMisc’* package was used to estimate Spearman’s correlation between the BA intensities in the stool, community BA exchange reaction potentials, and related microbial abundances. The p-values were adjusted for FDR at (p.adjusted < 0.05). Results are plotted using *‘heatmap*.*2’* function of *‘gplots’* package (v.3.0.4).

## Data Availability

Metagenomic sequencing data can be downloaded from
https://diabimmune.broadinstitute.org/diabimmune/ (NCBI BioProject ID: PRJNA231909).
The targeted bile acid metabolomics datasets generated in this study will be submitted to the NIH Common Fund’s National Metabolomics Data Repository (NMDR) website, the Metabolomics Workbench (https://www.metabolomicsworkbench.org).
Scripts for GSMM and data analysis can be downloaded from:
https://github.com/parthoBTK/GSMM_T1D.git

## Abbreviations

AED: average European diet
AGORA: assembly of gut organisms through reconstruction and analysis
BAs: bile acids
BSH: bile salt hydrolase
CA: cholic acid
CDCA: chenodeoxycholic acid
FSP: faecal secretion potential
EMP: estimated metabolic potential
FXR: farnesoid X receptor
GCA: glycocholic acid
GCDCA: glycochenodeoxycholic acid
GDCA: glycodeoxycholic acid
GEMs: genome-scale metabolic models
GSMM: genome-scale metabolic modeling
HCA: hyocholic acid
MCA: muricholic acids
MS: mass spectrometry
sIECs: small intestinal epithelial cells
TCA: taurocholic acid
TDCA: taurodeoxycholic acid
TGR5: Takeda G protein-coupled membrane receptor 5
THCA: taurohyocholic acid
UDCA: ursodeoxycholic acid
DCA: deoxycholic acid
UPLC-QqQMS: ultra-performance liquid chromatography-tandem mass-spectrometry
VMH: virtual metabolic human database

## Acknowledgements

This study was supported by the Academy of Finland postdoctoral grant (No. 323171 to S.L.), the Novo Nordisk Foundation (Grant no. NNF20OC0063971 to T.H. and M.O.), the Academy of Finland Centre of Excellence in Molecular Systems Immunology and Physiology Research 2012-17, No. 250114 to M.K. and M.O.), and the Academy of Finland project grant (No. 333981 to M.O.). Further support was received from the Swedish Research Council (grant no. 2016-05176 to T.H and M.O), Formas (grant no. 2019-00869 to T.H and M.O) and the Medical Research Funds, Helsinki University Hospital (to M.K.).

The authors thank Tuomas Mikael Lindeman and Peppi Raunioniemi (University of Turku) for excellent technical support in bile acid analysis, and to Dr. Aidan McGlinchey (Örebro University) for editing.

## Author contributions

Conceptualization (S.L., P.S., M.O.); Data curation (S.L., P.S., M.A.A.); Formal analysis (S.L., P.S.), Funding acquisition (S.L., T.H., M.K., M.O.); Investigation (S.L., P.S., T.H., J.H., M.K., M.O.); Methodology (P.S., A.M.D., M.A.A., T.H., M.O.); Resources (T.V., R.J.X., T.H., M.K.); Supervision (T.-H., M.K., M.O.); Roles/Writing - first draft (S.L., P.S., M.O.); Writing – critical review & editing (all authors).

## Data and code availability

Metagenomic sequencing data can be downloaded from https://diabimmune.broadinstitute.org/diabimmune/ (NCBI BioProject ID: PRJNA231909).

The targeted bile acid metabolomics datasets generated in this study will be submitted to the NIH Common Fund’s National Metabolomics Data Repository (NMDR) website, the Metabolomics Workbench (https://www.metabolomicsworkbench.org).

Scripts for GSMM and data analysis can be downloaded from: https://github.com/parthoBTK/GSMM_T1D.git

## Supplementary Notes

Lamichhane *et al*.,

**Figure S1.**
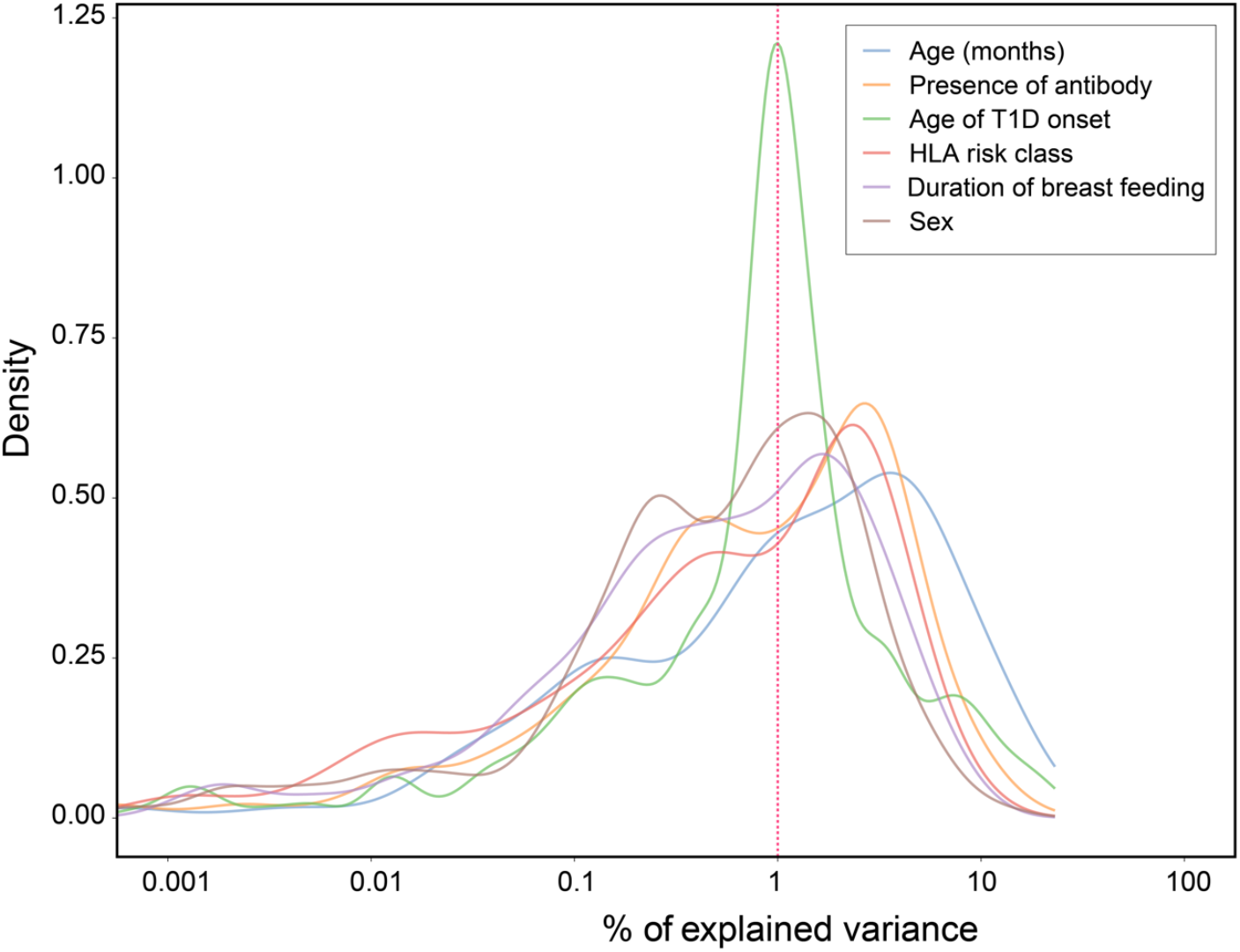
Factor analysis and identification of confounding factors affecting the microbiome. A density plot showing sample-wide distribution of (% of explained variances (EVs)) of various clinical and demographic factors associated with the normalized metagenomic analyzed in (n=111) stool samples.

**Figure S2.**
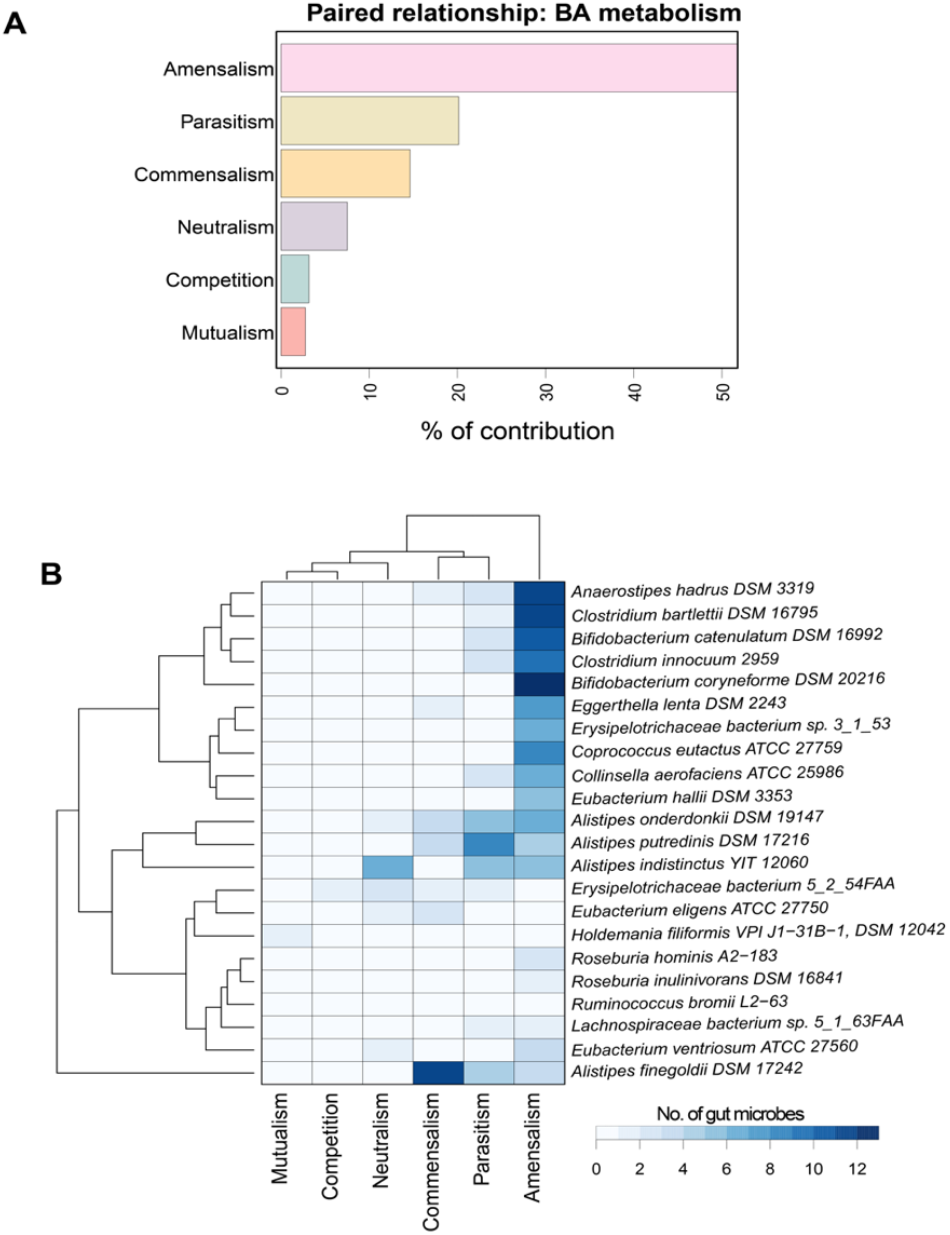
Modeling of pairwise interactions between 25 selected gut bacterial strains (*co-growth*) subjected. **(A)** The predicted relationship between all possible pairs of examined bacteria engaged in BA de-conjugation and BA biotransformation. *Amensalism*: In the paired simulation, one microbe grows slower than the other does, but their growth is not mutually dependent. *Parasitism*: One microbe grows faster than another does. *Competition*: In the paired simulation, both microbes exhibit slower growth as compared with their individual growth. *Neutralism*: Independent and unaffected growth of the microbes in the paired simulations. *Commensalism*: Growth of one microbe is faster and unaffected by the other. The definitions of mentioned concepts are derived from (Magnusdottir *et al*., 2017). **(B)** Gut bacterial strain pairs that can biotransform BAs were grouped by their genus.

**Figure S3.**
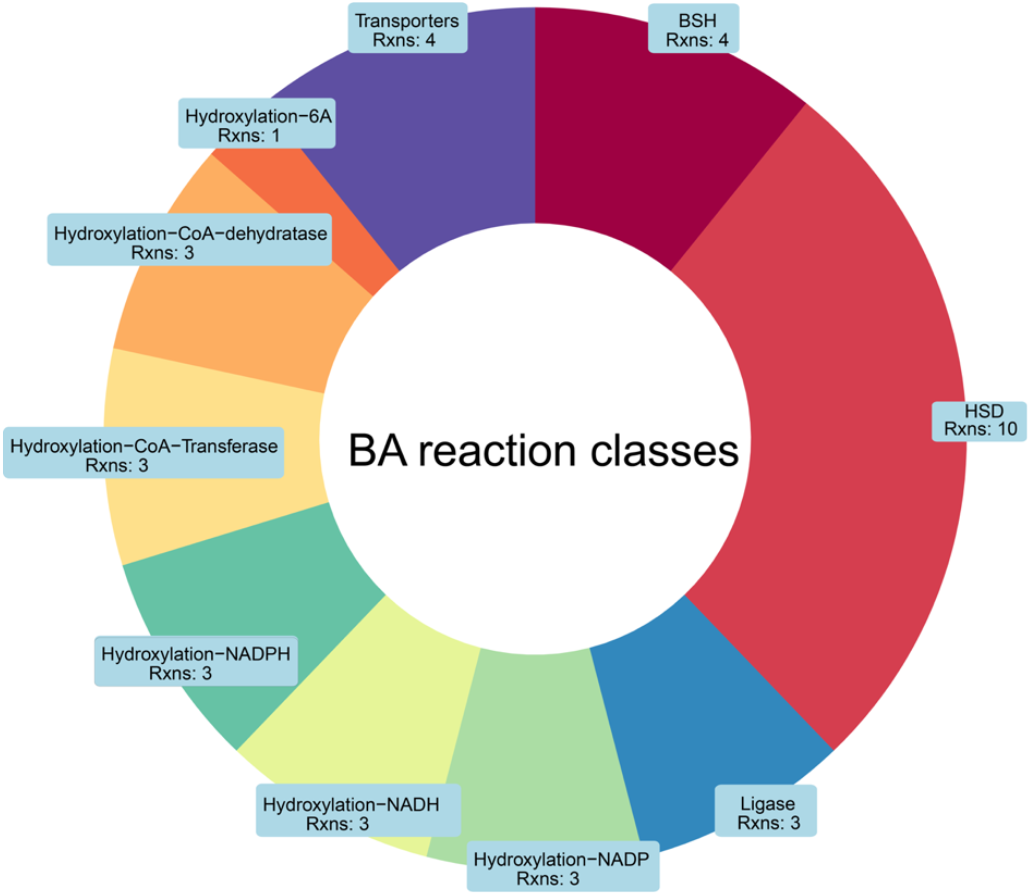
Bile acid reaction classes exhibited by the 25 abundant microbes. The BA reactions/pathways exhibited by human gut microbes spans between 10 different reaction classes that can carry out deconjugation, dehydrogenation, dehydroxylation and epimerization of BAs in the gut.

**Table S1.** Multivariate associations using linear models were performed with co-variates including age, sex, and case status (P2Ab or P1Ab or CTRs) in stool.

| Metadata | Features | coefficient | P value | Q value |
| --- | --- | --- | --- | --- |
| Age | LCA | 1.765868 | 3.19E-35 | 3.95E-33 |
| Age | HCA | -0.71767 | 1.96E-23 | 1.21E-21 |
| Age | CA | -1.01563 | 2.19E-20 | 9.04E-19 |
| Age | CDCA | -0.9442 | 3.85E-19 | 1.01E-17 |
| Age | HDCA | 1.017782 | 4.07E-19 | 1.01E-17 |
| Age | GLCA | 0.488118 | 5.37E-19 | 1.11E-17 |
| Age | GDHCA | -0.18543 | 1.16E-15 | 2.05E-14 |
| Age | TDCA | 0.451116 | 1.21E-10 | 1.88E-09 |
| Age | OXODCA | -0.58323 | 2.75E-10 | 3.79E-09 |
| Age | UDCA | 0.807496 | 1.29E-09 | 1.52E-08 |
| Age | GUDCA | 0.549565 | 1.35E-09 | 1.52E-08 |
| Age | DCA | -0.5483 | 7.77E-09 | 8.03E-08 |
| Age | THCA | -0.50224 | 8.87E-09 | 8.46E-08 |
| Age | TaMCA | -0.64066 | 1.05E-08 | 9.29E-08 |
| Age | GHDCA | 0.531877 | 4.10E-08 | 3.39E-07 |
| Age | bMCA | -0.5654 | 6.81E-08 | 5.28E-07 |
| Age | TLCA | 0.238253 | 5.38E-07 | 3.93E-06 |
| Age | oxoHDCA | -0.42934 | 4.05E-05 | 0.000279 |
| Age | GHCA | -0.1721 | 8.74E-05 | 0.00057 |
| Age | TDHCA | -0.16553 | 0.000214 | 0.001328 |
| Age | GCDCA | 0.203357 | 0.001128 | 0.006658 |
| Age | GDCA | 0.203259 | 0.001189 | 0.006701 |
| Age | aMCA | -0.1961 | 0.001382 | 0.007451 |
| Case | GUDCA | -0.66431 | 0.009749 | 0.050371 |
| Case | GHDCA | -0.6725 | 0.014903 | 0.073919 |
| Case | TUDCA | -0.7258 | 0.026109 | 0.11991 |
| Case | HDCA | -0.54519 | 0.028887 | 0.12793 |
| Case | THDCA | -0.65175 | 0.044398 | 0.189841 |
| sex | HDCA | -0.26414 | 0.017872 | 0.085236 |

**Table S2.** Multivariate associations using linear models were performed with co-variates including age, sex, and case status (P2Ab or P1Ab or CTRs) in serum.

| Metadata | Feature | Coefficient | P value | Q value |
| --- | --- | --- | --- | --- |
| Age | TDCA | 0.810951 | 4.84E-34 | 5.61E-32 |
| Age | HCA | -0.615 | 5.16E-29 | 2.99E-27 |
| Age | GLCA | 0.366212 | 3.35E-25 | 1.29E-23 |
| Age | TaMCA | -0.66722 | 7.47E-21 | 2.17E-19 |
| Age | GUDCA | 0.692191 | 2.46E-18 | 5.72E-17 |
| Age | THCA | -0.4358 | 5.13E-16 | 9.92E-15 |
| Age | UDCA | 0.584086 | 6.06E-15 | 1.00E-13 |
| Age | GHDCA | 0.650438 | 2.03E-13 | 2.95E-12 |
| Age | X7oxoHDCA | -0.33823 | 3.78E-11 | 4.88E-10 |
| Age | GDCA | 0.992672 | 7.50E-11 | 8.70E-10 |
| Age | TLCA | 0.16878 | 9.46E-10 | 9.98E-09 |
| Age | HDCA | 0.503645 | 4.96E-09 | 4.79E-08 |
| Age | X7OXODCA | -0.35812 | 6.31E-08 | 5.63E-07 |
| Age | TCA | -0.38511 | 1.41E-07 | 1.17E-06 |
| Age | TwMCA | 0.239079 | 1.54E-07 | 1.19E-06 |
| Age | TCDCA | -0.54483 | 3.05E-07 | 2.21E-06 |
| Age | CDCA | -0.28592 | 8.93E-07 | 6.10E-06 |
| Age | GHCA | -0.24791 | 1.23E-06 | 7.92E-06 |
| Age | TUDCA | 0.24461 | 1.94E-06 | 1.19E-05 |
| Age | THDCA | 0.235196 | 2.78E-06 | 1.61E-05 |
| Age | wMCA | -0.06713 | 0.007594 | 0.040039 |
| Case | HDCA | -0.5611 | 0.02448 | 0.118321 |
| sex | TDHCA | 0.019688 | 0.004911 | 0.027125 |
| sex | GHCA | 0.159941 | 0.013417 | 0.067668 |
| sex | GCA | 0.16621 | 0.034349 | 0.159378 |
| sex | THCA | 0.137237 | 0.04182 | 0.186583 |

**Table S3.** Multivariate associations using linear models were performed with co-variates including age, sex, and case status (P2Ab or P1Ab or CTRs) in the stool microbiome dataset.

| Metadata | feature | coef | pval | qval |
| --- | --- | --- | --- | --- |
| Age | t_GCF_000296465 | 0.004602 | 0.000156 | 0.07719 |
| Age | t_GCF_000154465 | 0.007676 | 0.000153 | 0.07719 |
| Age | t_Ruminococcus_gnavus_unclassified | -0.02405 | 0.000309 | 0.101766 |
| Age | t_GCF_000311925 | 0.001811 | 0.000427 | 0.105464 |
| Age | t_Bifidobacterium_breve_unclassified | -0.02191 | 0.000804 | 0.132415 |
| Age | t_GCF_000190535 | 0.002788 | 0.000776 | 0.132415 |
| Age | t_GCF_000146185 | 0.006975 | 0.00144 | 0.182298 |
| Age | t_GCF_000469345 | 0.000243 | 0.00169 | 0.182298 |
| Age | t_Veillonella_parvula_unclassified | -0.00865 | 0.001845 | 0.182298 |
| Age | t_Escherichia_coli_unclassified | -0.01997 | 0.002691 | 0.221572 |
| Age | t_Streptococcus_mitis_oralis_pneumoniae_unclassified | -9.06E-05 | 0.003431 | 0.260742 |
| Age | t_GCF_000265365 | 0.002121 | 0.006453 | 0.455398 |
| Age | t_Bacteroides_ovatus_unclassified | 0.018325 | 0.011665 | 0.638927 |
| Age | t_GCF_000153885 | 0.000159 | 0.012934 | 0.638927 |
| Age | t_GCF_000209875 | 0.008028 | 0.012914 | 0.638927 |
| Age | t_GCF_000468015 | 0.000374 | 0.011642 | 0.638927 |
| Age | t_GCF_000185705 | 3.22E-05 | 0.010589 | 0.638927 |
| Case | t_GCF_000159975 | 0.009565 | 0.001704 | 0.182298 |
| Case | t_GCF_000157995 | 0.000447 | 0.002104 | 0.188982 |
| Case | t_GCF_000218385 | 0.000305 | 0.010119 | 0.638927 |
| Case | t_GCF_000190535 | 0.006662 | 0.021796 | 0.800568 |
| Case | t_GCF_000242155 | -0.00049 | 0.022627 | 0.800568 |
| Case | t_GCF_000205025 | 0.00103 | 0.022923 | 0.800568 |
| Case | t_Lactobacillus_ruminis_unclassified | 0.002381 | 0.030614 | 0.800568 |
| Case | t_Faecalibacterium_prausnitzii_unclassified | 0.032737 | 0.035679 | 0.800568 |
| Case | t_GCF_000239295 | 0.001043 | 0.037556 | 0.800568 |
| Case | t_Bacteroides_ovatus_unclassified | 0.042377 | 0.039878 | 0.800568 |
| Case | t_GCF_000169035 | -0.0067 | 0.042992 | 0.800568 |
| Case | t_Streptococcus_parasanguinis_unclassified | 0.000385 | 0.048034 | 0.800568 |
| sex | t_Lactococcus_lactis_unclassified | 0.000553 | 0.037624 | 0.800568 |
| sex | t_Streptococcus_mitis_oralis_pneumoniae_unclassified | -6.30E-05 | 0.039398 | 0.800568 |
| sex | t_Clostridium_symbiosum_unclassified | 0.000892 | 0.046542 | 0.800568 |
| sex | t_GCF_000157995 | 0.000111 | 0.047607 | 0.800568 |

